# Taste alteration in COVID-19: a rapid review with data synthesis reveals significant geographical differences

**DOI:** 10.1101/2020.09.11.20192831

**Authors:** Nicola Cirillo

## Abstract

To facilitate a timely understanding of the differences in the prevalence of gustatory disturbances (GD) in individuals infected with SARS-CoV-2, we undertook a rapid systematic review of articles published in the repository of the National Library of Medicine (MEDLINE/PubMed) and medRxiv from their inception until September 3, 2020. The minimum requirements for completing a restricted systematic review were met. Of the 431 articles retrieved, 61 eligible studies (28,374 confirmed COVID-19 cases) from 20 countries were included in the analysis. The results show strong significant differences in the overall reported prevalence of GD between East Asia [13%, 95% CI 0.14-26.06%], Middle East [38.83%, 95% CI 27.47-50.19%], Europe [57.18%, 95% CI 52.35-62.01%], and The Americas [66.78%, 95% CI 54.77-78.79%]. There were no trends showing differences of GD prevalence in the available literature between February and August, 2020. These data show that there is a distinct geographical distribution of GD in COVID-19 patients and this may explain the differences of diagnostic criteria for COVID-19 case definition.

## Introduction

Early detection is key to reduce the spread of COVID-19 and is particularly challenging for asymptomatic or paucisymptomatic patients. Sudden loss of taste (ageusia), with or without loss of smell (anosmia) has been increasingly cited as independent symptoms or in association with the most common manifestations of the disease, such as fever, cough and fatigue (Guan et al., 2020). Unlike early studies (Lovato and de Filippis, 2020; O’Donovan et al., 2020), recent systematic reviews have shown that gustatory dysfunctions including ageusia, hypogeusia, and dysgeusia have a pooled prevalence of 81.6% of COVID-19 patients (Passarelli et al., 2020). Furthermore, loss of taste and smell have been reported to be distinguishing symptoms of COVID-19 with a high predictive value (Dawson et al., 2020). The European Centre for Disease Prevention and Control (EDCD) was one of the first public health bodies to include sudden onset of anosmia, ageusia or dysgeusia as chief clinical criteria to identify possible COVID-19 cases (ECDC, 2020). However, these symptoms have not been unanimously used for case identification and testing prioritization until August 2020. The Centers for Disease Control and Prevention (CDC) in the USA has updated COVID-19 case definition on 5 August to include new taste disorders as a chief clinical criterion for diagnosis (CDC, 2020). Soon afterwards (7 August), the World Health Organization (WHO) COVID-19 case definition was updated and included recent onset of ageusia, in the absence of any other identified cause, as suggestive of a probable COVID-19 case (WHO, 2020).

We hypothesized that differences in the criteria used for case identification of national or local public health bodies may reflect, at least in part, the changes of known prevalence rates of these symptoms over time and in a geographically specific manner. Because of the importance of the individuation of these trends in a timely fashion, we undertook a rapid systematic review with the aim of gathering overall data on the prevalence of taste alterations worldwide and in four large geographical areas including East Asia, Middle East (including Turkey), Europe (including Britain), and the Americas.

## Methods

### Study design and literature search

This study was conducted in accordance with the Preferred Reporting Items for Systematic Reviews and Meta-analyses (PRISMA) guidelines and used a rapid review approach due to time constraints (Khangura et al., 2012). The minimum requirements for completing a restricted systematic review (Plüddemann et al., 2018) were met. Accordingly, the search was performed by one investigator (N.C.) and verification of random samples off full texts for the accuracy of title/abstract screening and data extraction was undertaken by the same reviewer. Key terms used for the search were (SARS-CoV-2 or COVID or COVID-19) AND (taste or ageusia or hypogeusia or dysgeusia or gustatory). The search was conducted in PubMed/MEDLINE as well as medRxiv using advanced search (title and abstract) tool.

### Study selection and data extraction

The exclusion criteria were as follows: articles not in English, duplicate publications, irrelevant articles, studies where the infection status was not clearly confirmed, studies that did not evaluate gustatory outcomes individually, simple case reports, and review or systematic review articles. Studies using telephone surveys or Apps were only included where the respondents had a confirmed COVID-19 diagnosis. For studies reporting on cases from two or more geographical areas (e.g. East Asia and Europe), data for subgroup analysis was extracted only when information from individual countries was available. Where end date of patient recruitment was not provided, date of article submission was used as a surrogate information. The primary outcome was to assess the prevalence of gustatory alterations (ageusia, hypogeusia, dysgeusia) in confirmed COVID-19 cases worldwide and in geographical areas, whereas the secondary outcome was to establish a spatiotemporal pattern of published cases. No constraints were placed on the size of the cohorts to ensure a comprehensive search and to identify the maximum number of potential articles.

### Statistical analysis

Subgroup analyses were performed based on the country of origin of the studies by pooling the actual data reported in each individual study. Difference in prevalence and categories among subgroups was assessed with chi-square statistics and one-way ANOVA, as appropriate. Tukey’s post-hoc test or Student’s t test were used for comparison between group pairs. By making a further assumption that the dependent variable may not be normally distributed, the Kruskal-Wallis test was also used to compare overall differences in prevalence. Where appropriate, Pearson’s coefficient was used to assess the correlation between time and prevalence. A level of p <0.05 was chosen to determine statistical significance.

## Results

Our search identified 431 studies and full text of 91 relevant articles meeting the inclusion criteria were assessed. Of these, 61 studies were included in the data synthesis (**Figure 1**). Studies were from 20 different countries, and 5 derived from multi-national collaborations. Most cohorts were from Europe (n=40), followed by Middle East (n=8), North and South America (n=6 and n=2, respectively), East Asia (n=6) and Africa (n=1). Two articles (Lechien et al., 2020a; 2020b) pooled multinational data within Europe, whereas three studies (Chiesa-Estomba et al., 2020a; Menni et al., 2020; Qiu et al., 2020) included cases from two main geographical areas. Study populations and prevalence range are graphically depicted in **Figure 2**.

**Figure 1.**
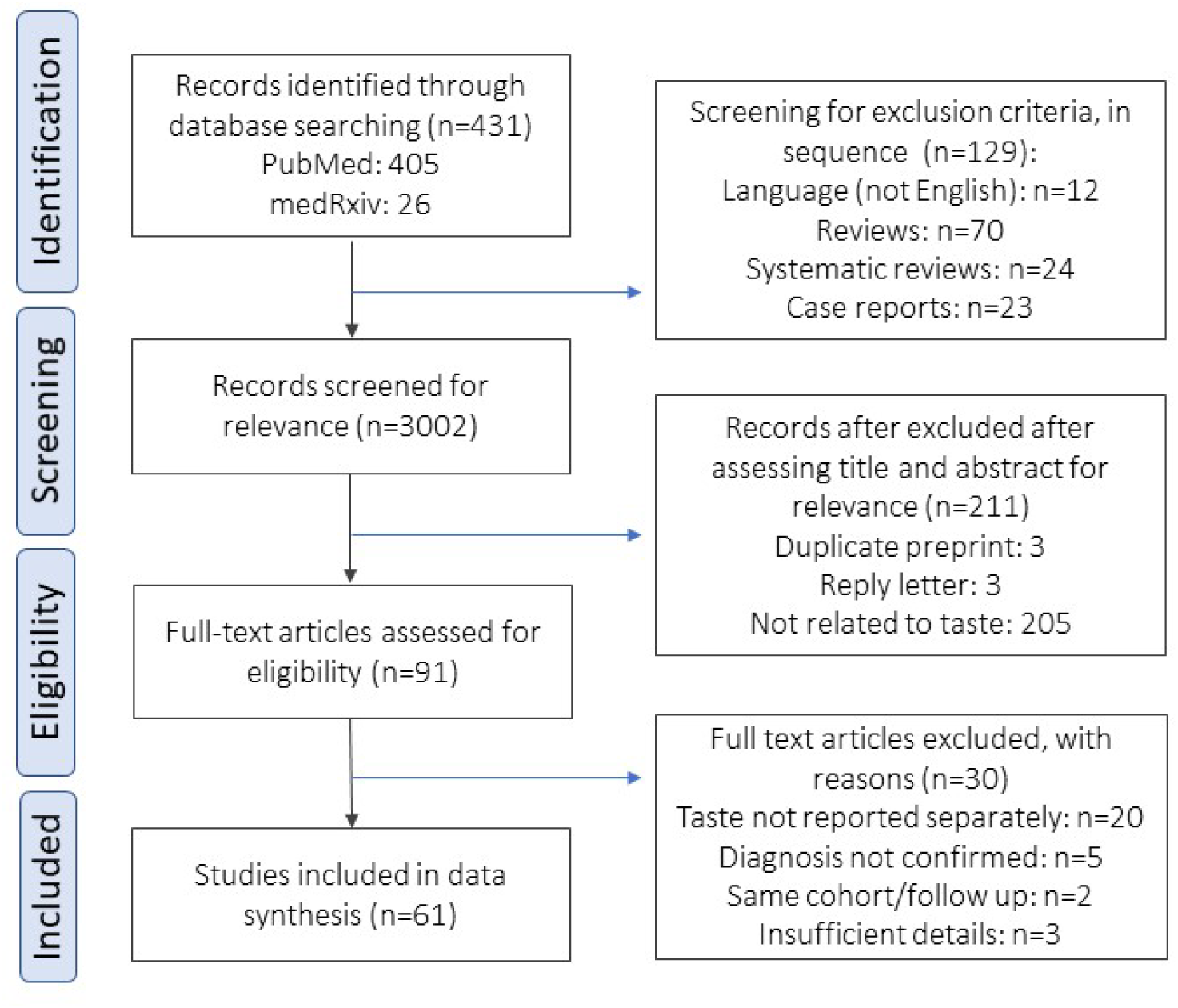
Preferred Reporting Items for Systematic Reviews and Meta-Analyses (PRISMA) flow chart of study selection process.

**Figure 2.**
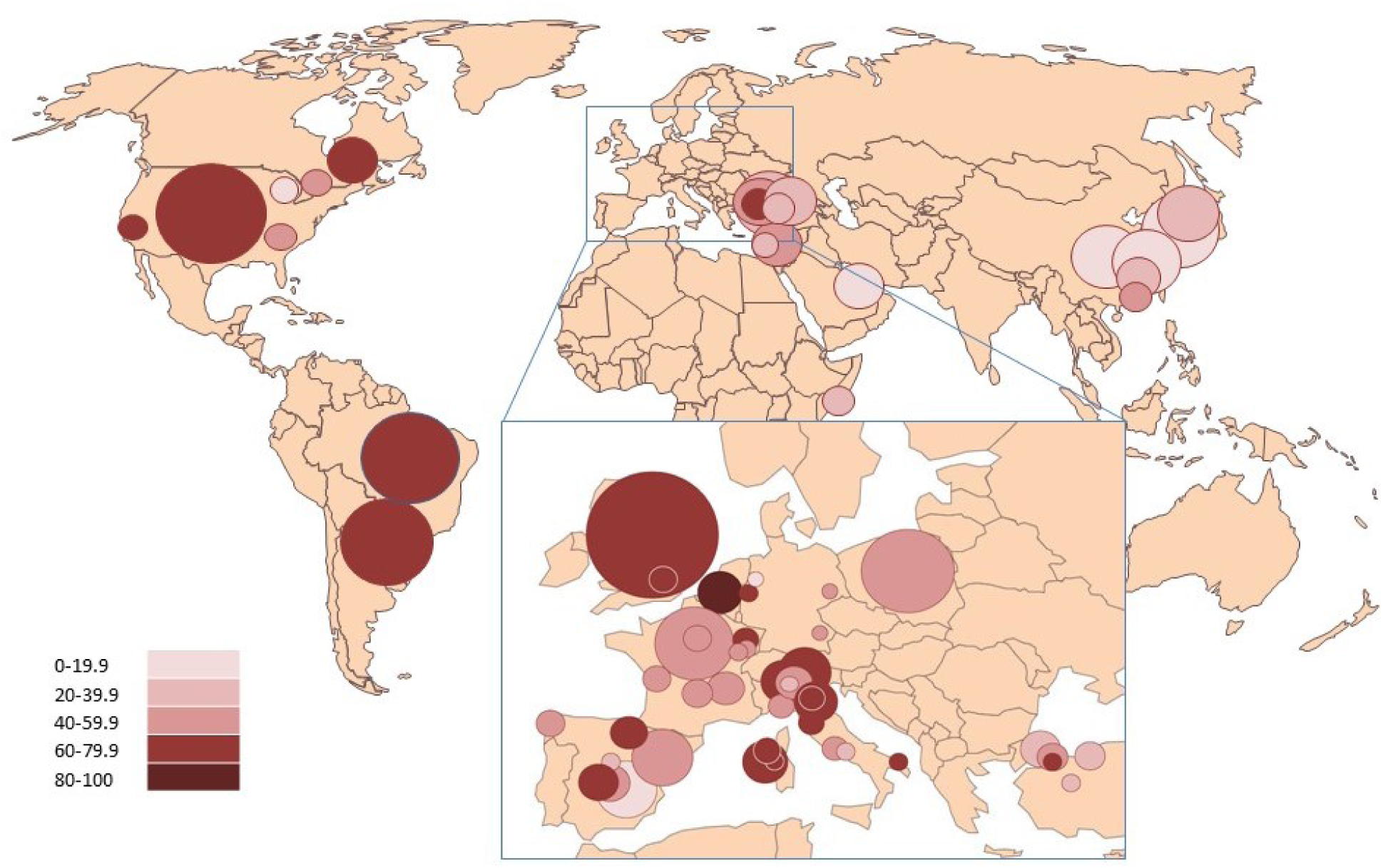
Worldwide prevalence of gustatory disorders (GD) in COVID-19 positive cases. Circle size is proportional to study population.

Worldwide, 14,486 out of 28,374 confirmed COVID-19 cases (51.05%) reported subjective and/or objective GD (**Table 1**). Strikingly, there were significant differences of prevalence between subgroups with either of the assumptions (ANOVA, p < 0.000106; Kruskal-Wallis, p < 0.00071). When comparing each geographical region, there were significant differences between all subgroups except Europe vs America (**Suppl Table 1**). Studies from East Asia reported the lowest prevalence of GD (13%, 95% CI 0.14-26.06%), followed by Middle East (38.83%, 95% CI 27.47-50.19%), Europe (57.18%, 95% CI 52.35-62.01%), and The Americas (66.78%, 95% CI 54.77-78.79). We could not find trends in differences in gross GD prevalence in COVID-19 patients over time, except in East Asia (**Supplementary Figure 1**).

**Table 1.** Studies included for the evaluation of gustatory dysfunction (GD).

| Reference | GD | COVID+ | Prevalence | Area |
| --- | --- | --- | --- | --- |
| Romero-Sanchez et al, 2020 | 52 | 841 | 6.2% | Spain |
| Paderno et al., 2020 | 320 | 508 | 63% | Italy |
| Zayet et al., 2020b | 62 | 95 | 65% | France |
| Vaira et al., 2020a | 39 | 72 | 54.2% | Italy |
| Lapostolle et al., 2020 | 757 | 1487 | 50.9% | France |
| Martin-Sanz et al., 2020 | 114 | 215 | 53% | Spain |
| Lechien et al., 2020c | 770 | 1420 | 54.2% | Europe (multiple sites) |
| Villarreal et al., 2020 | 161 | 230 | 70% | Spain |
| Petrocelli et al., 2020 | 184 | 300 | 61.3% | Italy |
| Qiu C et al., 2020 | 50, 20 | 116, 39 | 45.16% | France, Germany (and China) |
| Abalo-Lojo et al., 2020 | 74 | 131 | 56.5% | Spain |
| Zayet et al., 2020a | 34 | 70 | 48.6% | France |
| Tudrej et al., 2020 | 92 | 198 | 46.5% | France |
| Poncet-Megemont et al., 2020 | 81 | 139 | 58.3% | France |
| Patel et al., 2020 | 89 | 141 | 63.1% | UK |
| Chary et al., 2020 | 64 | 115 | 55.65% | France |
| Sierpinski et al., 2020 | 923 | 1942 | 47.5% | Poland |
| Izquierdo-Dominguez et al., 2020 | 442 | 846 | 52.2% | Spain |
| Rojas-Lechuga et al., 2020 | 128 | 197 | 65% | Spain |
| Giacomelli et al., 2020 | 17 | 59 | 28.8% | Italy |
| Menni et al., 2020 | 4178 | 6452 | 64.76% | UK (and USA) |
| Vaira et al., 2020b | 76 | 106 | 71.7% | Italy |
| Mercante et al., 2020 | 113 | 204 | 55.4% | Italy |
| Lechien et al., 2020b | 1136 | 2013 | 56% | Europe (multiple sites) |
| Klopfenstein et al., 2020 | 34 | 70 | 48% | France |
| Liguori et al., 2020 | 48 | 103 | 46.6% | Italy |
| Luers et al., 2020 | 50 | 72 | 69.4% | Germany |
| Dell'Era et al., 2020 | 232 | 355 | 65.4% | Italy |
| Magnavita et al., 2020 | 31 | 82 | 37.8% | Italy |
| Meini et al., 2020 | 69 | 100 | 69% | Italy |
| Lechien et al., 2020a | 342 | 385 | 88.8% | Belgium |
| Beltrán-Corbellini et al, 2020 | 28 | 79 | 35.44% | Spain |
| Vaira et al., 2020d | 234 | 340 | 67.8% | Italy |
| Gelardi et al., 2020 | 52 | 72 | 72.2% | Italy |
| Vacchiano et al., 2020 | 66 | 108 | 61% | Italy |
| De Maria et al., 2020 | 48 | 95 | 50.5% | Italy |
| Fistera et al., 2020 | 6 | 43 | 14% | Germany |
| Vaira et al., 2020c | 91 | 138 | 65.9% | Italy |
| Hintschich et al., 2020 | 18 | 41 | 44% | Germany |
| Chiesa-Estomba et al., 2020a | 718 | 1043 | 68.8% | Europe (multiple sites) |
|  | **12,043** | **21,062** | **57.18%** | **TOTAL EUROPE** |
| Yan et al., 2020 | 42 | 59 | 71% | USA |
| Menni et al., 2020 | 490 | 726 | 67.49% | USA (and UK) |
| Carignan et al., 2020 | 85 | 134 | 63.4% | Canada |
| Pinna et al., 2020 | 10 | 50 | 10% | USA |
| Chiesa-Estomba et al., 2020 | 333 | 542 | 61.4% | South America<br>(Multiple sites) |
| Lee et al., 2020 | 32 | 56 | 57.1% | Canada |
| Kempker et al., 2020 | 27 | 51 | 52.9% | USA |
| Brandao Neto et al., 2020 | 499 | 655 | 76.2% | Brazil |
|  | **1518** | **2273** | **66.78%** | **TOTAL AMERICA** |
| Sayin et al., 2020 | 46 | 64 | 71.9% | Turkey |
| Biadsee et al., 2020 | 67 | 128 | 52% | Israel |
| Altin et al., 2020 | 22 | 81 | 27.2% | Turkey |
| Salepci et al., 2020 | 77 | 223 | 34.5% | Turkey |
| Al-Ani and Acharya, 2020 | 28 | 141 | 19.86% | Qatar |
| Sakalli et al., 2020 | 81 | 172 | 47.1% | Turkey |
| Levinson et al., 2020 | 14 | 42 | 33.3% | Israel |
| Çalica Utku et al., 2020 | 51 | 143 | 35.7% | Turkey |
|  | **386** | **994** | **38.83%** | **TOTAL MIDDLE EAST** |
| Lee et al., 2020 | 353 | 3191 | 11% | Korea |
| Mao et al., 2020 | 12 | 214 | 5.6% | China |
| Kim et al., 2020 | 58 | 172 | 33.7% | Korea |
| Qiu et al., 2020 | 30 | 239 | 12.55% | China (and France,<br>Germany) |
| Cho et al., 2020 | 36 | 83 | 43.4% | Hong Kong |
| Liang et al., 2020 | 33 | 86 | 38.4% | China |
|  | **522** | **3985** | **13.1%** | **TOTAL EAST ASIA** |
| Farah YMM et al., 2020 | 17 | 60 | 28.3% | Somalia |
|  | **14,486** | **28,374** | **50.05%** | **WORLD** |

## Discussion

Awareness of the association between taste alterations and COVID-19 could be key for diagnosing the disease, particularly in dental and oral health settings (Cirillo, 2020). We followed a streamlined approach to synthesizing evidence, the rapid review, which is typically used for informing emergent decisions faced by decision makers in health care settings (Khangura et al., 2012). The results of this rapid systematic review show that there are distinct geographical patterns of GD in patients with established SARS-CoV-2 infection.

The first systematic assessments of the evidence available up to March 2020 failed to identify associations between anosmia/ageusia and COVID-19 (Lovato and de Filippis, 2020; O’Donovan et al., 2020). For example, of the studies included in an early systematic review (with a total of 1556 patients), none reported about olfactory or gustative dysfunctions (Lovato and de Filippis, 2020). When looking specifically at the evidence for anosmia in COVID-19 up to 23 March, researchers found it to be “limited and inconclusive” (O’Donovan et al., 2020). The first study reporting a 5.1 and 5.6% prevalence of hyposmia and hypogeusia, respectively, was a pre-print (non-peer-reviewed) case series of a Chinese population (Mao et al., 2020). In sharp contrast, a most recent meta-analysis analyzing smell and taste alterations not only reported that almost half of COVID-19 patients had these symptoms but, also, that 15% of patients had olfactory and gustatory abnormalities as their initial symptoms (Chi et al., 2020).

Recent systematic reviews assessing chemosensory alterations in COVID-19 patients have been published (Abdullahi et al., 2020; Almqvist et al., 2020; Agyeman et al., 2020; von Bartheld et al., 2020; Borsetto et al., 2020; Carrillo-Larco et al., 2020; Chen et al., 2020; Chi et al., 2020; da Costa et al., 2020; Hoang et al., 2020; Passarelli et al., 2020; Printza et al., 2020; Romoli et al., 2020; Samaranayake et al., 2020; Struyf et al., 2020; Tong et al., 2020; Wang et al., 2020; Whittaker et al., 2020), however only one review was specifically focused on taste changes (Aziz et al., 2020). In their pooled analysis, Aziz et al. (2020) found that almost half of the patients (49.8%) with COVID-19 have altered taste sensation. In the reviews assessing chemosensory alterations, the range of olfactory and gustatory alterations, when reported individually, was 3.2-100% and 0-92.6%, respectively. By and large, when the prevalence was pooled on the total number of cases examined, olfactory and gustatory alterations were found in approximately half of COVID-19 patients. Our data are in agreement with the existing literature on the worldwide incidence of GD: out of 28,374 confirmed COVID-19 cases, 14,486 (51.05%) reported GD. Overall, analysis of high-level evidence (Cirillo and Colella, unpublished) supports the inclusion of gustatory alterations as cardinal symptoms of COVID-19.

With regards to possible bias, it is to note that the majority of the studies are cross-sectional, retrospective observational studies, hence recollection bias may be present. Most studies are similar to those previously graded as “moderate risk of bias” (Tong et al., 2020). Importantly, the presence of taste alterations may not be reported in the presence of other severe symptoms, such as dyspnea, fever, and productive cough. This might explain the lack of association between GD and COVID-19 in the first studies published in February and March 2020 and the significant association with mild cases. For these reasons, the true prevalence of ageusia, hypogeusia and dysgeusia might be significantly higher than reported (Aziz et al., 2020).

Other potential weaknesses are that measures for assessing GD were not validated and that the definition of dysgeusia is not unanimously accepted. Furthermore, the awareness of the occurrence of taste alterations in COVID-19 cases may have determined and increase of self-reported GD over time (Menni et al., 2020), and/or this symptom may have been increasingly investigated by doctors when taking history of suspected cases. Although the latter remains a possibility, our data failed to show a trend of increased GD prevalence over time, except in East Asia. Unlike other studies (von Bartheld et al., 2020) our analysis focused on the geographical location of the studies rather than ethnicity. Although this approach may fail to individuate genetic/ethnic determinants of infection, we believe that our methodology is better suited to study the clinical manifestations of the disease and to inform the decisions of public health surveillance bodies.

Most studies relied on the individuals reporting about their subjective (self-reported) impressions, whereas a small number of studies used structured (objective) tests to assess taste dysfunction, for example by using substances with the four basic tastes (sweet, sour, salty, bitter) sprayed on the tongue in a supra-threshold dose. When a comparison between subjective and structured (objective) gustatory function was made, no significant differences were found (Agyeman et al., 2020). Therefore, self-reported taste alterations can be considered a reliable parameter for investigating the prevalence of this condition in COVID-19 patients.

## Conclusion

GD in COVID-19 show distinct geographical patterns. Given the potential usefulness of taste assessment in the diagnosis of mildly and paucisymptomatic patients, it is imperative to recognize ageusia/hypogeusia/dysgeusia as a potential clinical manifestation of COVID-19, particularly in Europe and America. Dentists may be the first healthcare providers to diagnose taste disturbances and are likely to play an important role in case identification and early diagnosis of COVID-19 cases in the near future.

## Data Availability

Full raw data available upon request

## Acknowledgements

N.C. would like to acknowledge the support of the Melbourne Dental School, The University of Melbourne.

## Supplementary Material

**Supplementary Figure 1.**
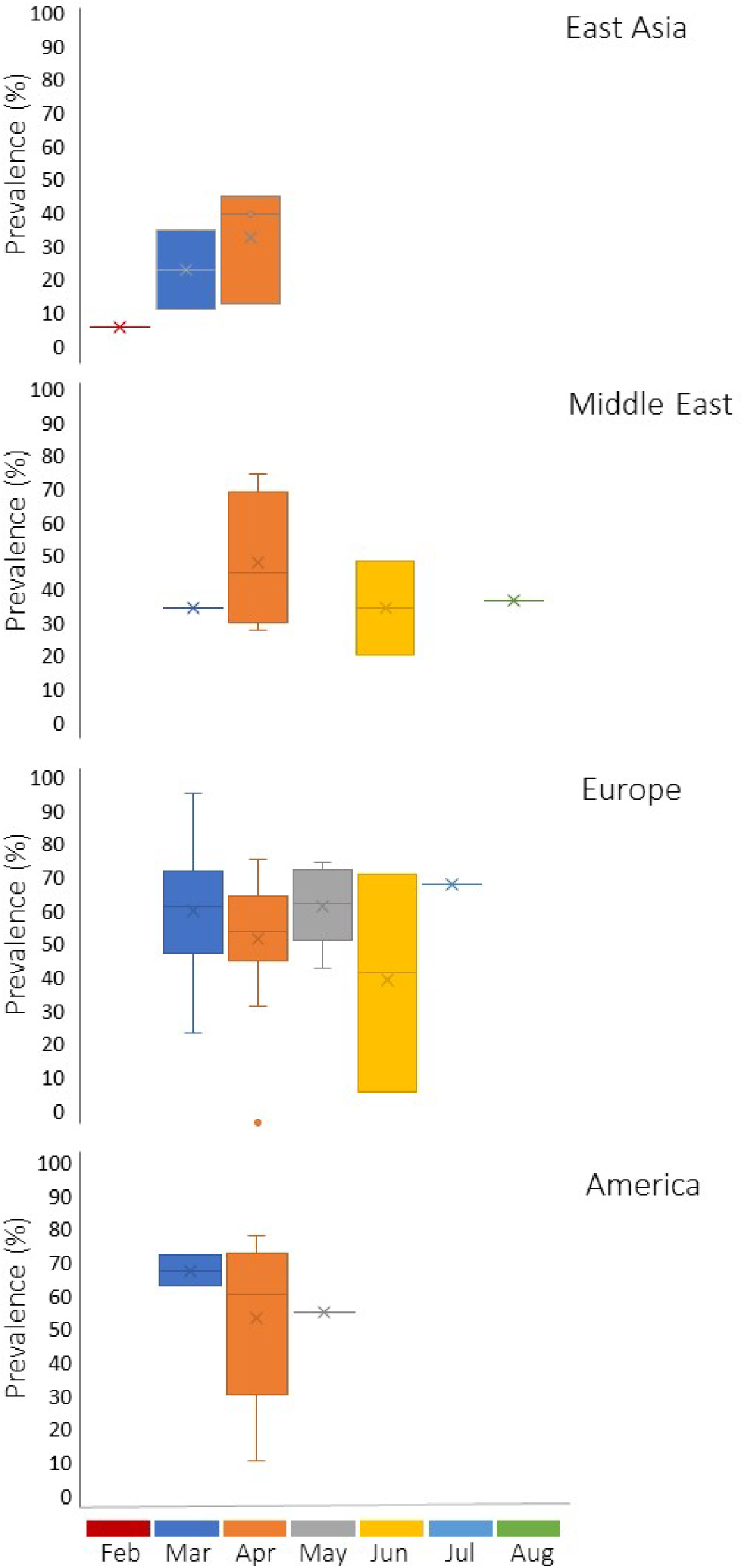
Reported prevalence (%) of GD in COVID-19 patients from February to August, 2020. End date of recruitment or date of submission was used to identify a temporal category.

**Supplementary Table S1.** *p* value of student’s t test comparing subgroups

|  | East Asia | Middle East | Europe | America |
| --- | --- | --- | --- | --- |
| East Asia |  | 0.047 | 0.002 | 0.001 |
| Middle East | 0.047 |  | 0.02 | 0.023 |
| Europe | 0.002 | 0.02 |  | 0.29 |
| America | 0.001 | 0.023 | 0.29 |  |

